# Pregnancy as a Diagnostic Catch-up Window in U.S. Health Claims

**DOI:** 10.64898/2026.11.23.26368725

**Authors:** Joseph Bommarito, Elizabeth Anderson, Lilly Springer, Yurong Zhang, Misty Heggeness

**Affiliations:** Institute for Policy and Social Research, University of Kansas, Lawrence, KS, USA; Department of Sociology, University of Kansas, Lawrence, KS, USA; Department of Economics and Department of Health Studies, American University, Washington, DC, USA; Institute for Policy and Social Research and School of Public Affairs, University of Kansas, Lawrence, KS, USA

**Keywords:** pregnancy, prenatal care, chronic hypertension, diabetes, sickle cell trait, insurance claims

## Abstract

Pregnancy brings many women into regular contact with healthcare and creates opportunities to recognize conditions that need attention beyond childbirth. We asked whether chronic conditions, unrelated to pregnancy itself, first appear in health care records more often as women enter pregnancy and have increased contact with the health system. We matched 175,505 women admitted for delivery in 2023 to women in the same US claims cohort whose pregnancies began later and who were not pregnant at the time. We compared billed outpatient evaluation days and first records of chronic hypertension, non-gestational diabetes, and sickle cell trait. Outpatient evaluation days rose after pregnancy began, while they changed little in the comparison group. Before 20 weeks of gestation, there were 2.4 additional first hypertension records per 1,000 women entering pregnancy (95% confidence interval 1.6 to 3.1). The difference was smaller for diabetes and much larger for sickle cell trait. We show that pregnancy is a window for bringing existing conditions into the claims record.

## Introduction

Pregnancy brings many women who may not have a history of routine care into contact with the healthcare system, creating opportunities to detect previously undiagnosed conditions. Blood pressure measurements, glucose testing, and hemoglobin testing are all commonplace and can reveal many conditions that existed before pregnancy [1-5]. In data commonly used to study maternal health, such as insurance claims or medical records, the first documented diagnosis during these encounters can appear to mark disease onset even when the disease was present prior to pregnancy but undiagnosed. Distinguishing when a condition enters the record from when it began matters for how we interpret maternal morbidity and plan postpartum care. More contact can bring more opportunities to identify a condition, even if each encounter is no more likely to yield a diagnosis. In the Oregon Medicaid experiment, coverage increased diabetes diagnoses and treatment without a detectable change in the prevalence of elevated glycated hemoglobin during follow-up [6]. In a matched study of prenatal care, additional blood pressure measurements through remote monitoring were associated with more early chronic hypertension diagnoses, even though the matched groups had similar numbers of prenatal visits [7]. These studies motivate separating contact frequency from what clinicians measure and record.

We examine whether first records of chronic hypertension, non-gestational diabetes, and sickle cell trait become more common when women enter pregnancy. We compare women admitted for delivery in 2023 with women with similar characteristics who are not pregnant at that time. We measure changes in billed outpatient evaluations, compare first records before and after pregnancy begins, and describe the additional diagnoses first coded during the delivery admission.

We chose chronic hypertension because blood pressure is measured at routine prenatal visits, giving clinicians repeated opportunities to recognize elevated blood pressure [1]. We included non-gestational diabetes because early prenatal glucose testing can uncover diabetes that may have been present before pregnancy [2-4]. Sickle cell trait provides an additional test because it is inherited and present from birth [5]. Its first appearance in claims during pregnancy therefore reflects documentation of an existing trait, although the claims cannot establish whether it was previously unknown to the patient or clinician. These traits do not represent the full range of possible interpretations of this paper. Our findings suggest that a variety of diseases may be present but undiagnosed prior to pregnancy-related health contact, and routine prenatal contact can uncover them. We conclude that pregnancy acts as an important diagnostic catch-up window, especially for women with less interaction with the healthcare system.

These questions also have an equity dimension. Sickle cell trait is more prevalent among Black Americans, although it occurs across racial and ethnic groups [8]. A study of young African American adults found substantial uncertainty about personal trait status [9]. At the same time, research with Black women has linked racism and economic insecurity to delayed or forgone reproductive care [10]. Women in lower socioeconomic categories, many of whom rely on Medicaid or are uninsured, have more limited healthcare interactions than those with commercial insurance. National survey data document barriers such as costs, transportation, childcare, and difficulty obtaining time away from work or school [11]. The American Rescue Plan Act of 2021 and the Consolidated Appropriations Act of 2023, now enacted in 49 of the 50 states, give new mothers 12 months of postpartum healthcare coverage. This can lead to not only vital postpartum care but also the beginning of treatment for chronic conditions previously undiscovered. Today, prenatal contact offers an important opportunity to identify or document previously unrecognized health needs among women facing barriers to routine care.

## Materials and methods

### Data sources

We used de-identified US health claims extracts licensed from the HealthVerity health claims ecosystem. The primary extract was the 2021–2024 Longitudinal delivery cohort from the August 2026 release. The extract includes women who gave birth during 2021–2024 and their complete recorded claims before and after delivery. Only women with insurance are included in the sample, meaning this study undercounts uninsured women. This file includes open and closed medical claims, medical and pharmacy enrollment records from participating closed-claims sources, and service dates, diagnosis codes, and procedure codes. We used the delivery, enrollment, and diagnosis tables from that extract and scanned the corresponding procedure records for outpatient evaluations and tests.

### Cohort and calendar matching

We identified each woman’s first eligible observed delivery admission in the derived 2021–2024 cohort. For women with multiple deliveries during the period, we retained only the first eligible observed delivery. The main analysis included women aged 18–49 with a 2023 admission for delivery and a recorded gestational age of 22–42 weeks. Restricting the analysis to 2023 births ensures we have two years of complete health records before delivery. We estimated gestational day zero by subtracting the number of completed gestational weeks coded on the delivery admission from the admission date [12]. We required continuous recorded medical enrollment beginning 449 days before the estimated start of pregnancy and continuing through delivery discharge. This allowed for a longitudinal look-back period of about 15 months before pregnancy began to find evidence of disease treatment before pregnancy-related healthcare contact. This allows us to distinguish a new condition appearing in the diagnosis record from a condition actively being treated. One limitation is that we cannot distinguish newly diagnosed diseases from previously known diagnoses that were not actively being treated. However, this look-back period allows us to be confident that even if the patient knew about the diagnosis, they were not receiving active treatment for it, and any uptick in treatment is still a meaningful effect.

Comparison women had their first eligible delivery admission in 2024. This represents a better comparison than women who never had a child or are far removed from children, as they are women who might also be beginning medical preparations for child-rearing. We assigned each comparison woman her matched partner’s estimated pregnancy start date as the shared index date and required any pregnancy of the comparison woman to begin after the 20-week comparison window ended. Matching was one-to-one without replacement within the pregnancy index calendar month, delivery state, five age groups (18–24, 25–29, 30–34, 35–39, and 40–49), and recorded medical payer at index (commercial insurance or managed Medicaid). This analysis represents the portion of the cohort with a contemporaneous match, resulting in an analytical sample of 351,010 women split evenly into 175,505 pairs of treated (i.e., women who gave birth) and untreated (i.e., women not currently pregnant).

### Diagnoses and billed care

For the primary comparisons, we used International Classification of Diseases, Tenth Revision, Clinical Modification (ICD-10-CM) codes from billed health records throughout the observational window. We defined chronic hypertension by I10, I11, I12, I13, or I15 and non-gestational diabetes by E10, E11, or E13 in the primary extract. Absence of a diagnosis code during the lookback does not necessarily mean the woman was unaware of a disease being present, but it does mean there is no evidence that she was actively receiving treatment for the disease. Given that diabetes, hypertension, and sickle cell trait all require some form of management, it is likely that few observations in the sample represent women who had access to insurance to pay for treatment they needed but chose not to get it. This concern would be larger if our sample included women who lacked access to insurance during this period, which is one reason they are excluded from this sample. In sensitivity analyses, we also counted O10 and O11 as hypertension outcomes, and O24.0, O24.1, O24.3, and O24.8 as diabetes outcomes. Gestational hypertension and gestational diabetes codes alone did not qualify, as we are specifically looking for diseases unrelated to childbirth. We analyzed D57.3 separately for sickle cell trait. Professional guidance recommends offering hemoglobinopathy testing before pregnancy or at the initial prenatal visit when prior results are unavailable [5].

We counted distinct days with a billed office or outpatient evaluation, using procedure codes 99201–99205, 99211–99215, 99241–99245, 99381–99387, 99391–99397, 99441–99443, G0463 and T1015. We excluded procedure lines with place-of-service codes for urgent care, inpatient hospital, or emergency department. We counted separately dated prenatal service markers (0500F, 0501F and 0502F) and hemoglobin testing (83020 and 83021) procedures. The measures capture billable evidence of contact and testing [13].

### Statistical analysis

To assess changes in healthcare contact, we counted each woman’s billed outpatient evaluation days during the 12 weeks before and the first 12 weeks after the pregnancy start date. We calculated the change between these periods for each woman, then subtracted the comparison woman’s change from that of the woman entering pregnancy. We averaged these differences across pairs to estimate how much more outpatient contact changed among women entering pregnancy.

For each condition, we compared the proportion of women with a first qualifying diagnosis record during three windows: the 12 weeks before pregnancy, the first 12 weeks after pregnancy onset, and the first 20 weeks after pregnancy begins. The 20-week window included the first 12 weeks and acts as a sensitivity test. We excluded a pair from a condition’s analysis if either woman had a record of that condition during the 365-day diagnosis-history period preceding the pre-pregnancy observation window. For the two post-index comparisons, we additionally excluded pairs in which either woman had a qualifying record during the 12 weeks immediately before pregnancy onset. These restrictions ensured that both members of a pair remained eligible to have a first record at the start of the relevant comparison. We calculated risk differences by subtracting the comparison-group proportion from the pregnancy-group proportion and expressed these differences per 1,000 women.

To describe when changes in contact and documentation occurred, we plotted results in four-week intervals from 12 weeks before pregnancy through the first 20 weeks after. Outpatient evaluation days were expressed per 100 woman-weeks. First diagnosis records were expressed per 1,000 woman-weeks at risk, with each woman contributing time through her first qualifying record (including that day) or the end of the observation period.

We also examined whether contact and documentation patterns differed by payer at delivery. We classified pairs by the pregnancy-group woman’s medical enrollment payer at delivery discharge: commercial insurance or managed Medicaid. We retained her original matched comparison woman regardless of that woman’s payer at her own delivery. Pairs with other, multiple, or unknown delivery payer were excluded from the primary delivery-payer comparison. To describe contact before pregnancy, we reported mean billed outpatient evaluation days and the proportion with no evaluation during the 365-day diagnosis-history period and the 12 weeks immediately before pregnancy. We then estimated changes in contact and first-record risk within each payer group using the same definitions and eligibility rules as the main analysis.

We tested payer differences directly by subtracting each commercial estimate from its managed Medicaid counterpart. Because the payer groups comprised separate matched pairs, the standard error of this contrast was the square root of the sum of the two estimated variances. We calculated Holm-adjusted p-values for four principal payer contrasts: the outpatient contact difference in changes and the three condition-specific risk differences before 20 weeks.

Finally, we calculated the likelihood of continued treatment one year after the initial diagnosis. A diagnosis entering the record has little value if the condition does not receive treatment in the future. For each mother who receives a diagnosis, we therefore test, by payer type, the likelihood that she continues to receive treatment for that condition one year or more after discharge.

For the matched risk differences and changes in outpatient contact, we calculated standard errors from the observed variation in within-pair differences and constructed 95% confidence intervals using a normal approximation. These calculations accounted for pairing while treating different matched pairs as independent.

## Results

### More chronic conditions entered the record

Of 369,473 women entering pregnancy, we matched 175,505 to comparison women. Managed Medicaid accounted for 61% of pairs. In the 12 weeks before and after pregnancy, billed outpatient evaluation days rose from 1.03 to 1.85 per woman in the pregnancy group and from 0.99 to 1.02 among comparison women over the 12-week period. The difference in changes was 0.80 days per woman (95% confidence interval 0.78 to 0.81). The increase began after the first four gestational weeks (Fig 1A). As expected, this shows that at pregnancy onset, the expected number of outpatient days sharply increases compared to women who are not pregnant. This effect is likely a conservative estimate of the actual effect, as multiple prenatal appointments are often bundled into a single billing, which can hide some visits.

**Fig 1.**
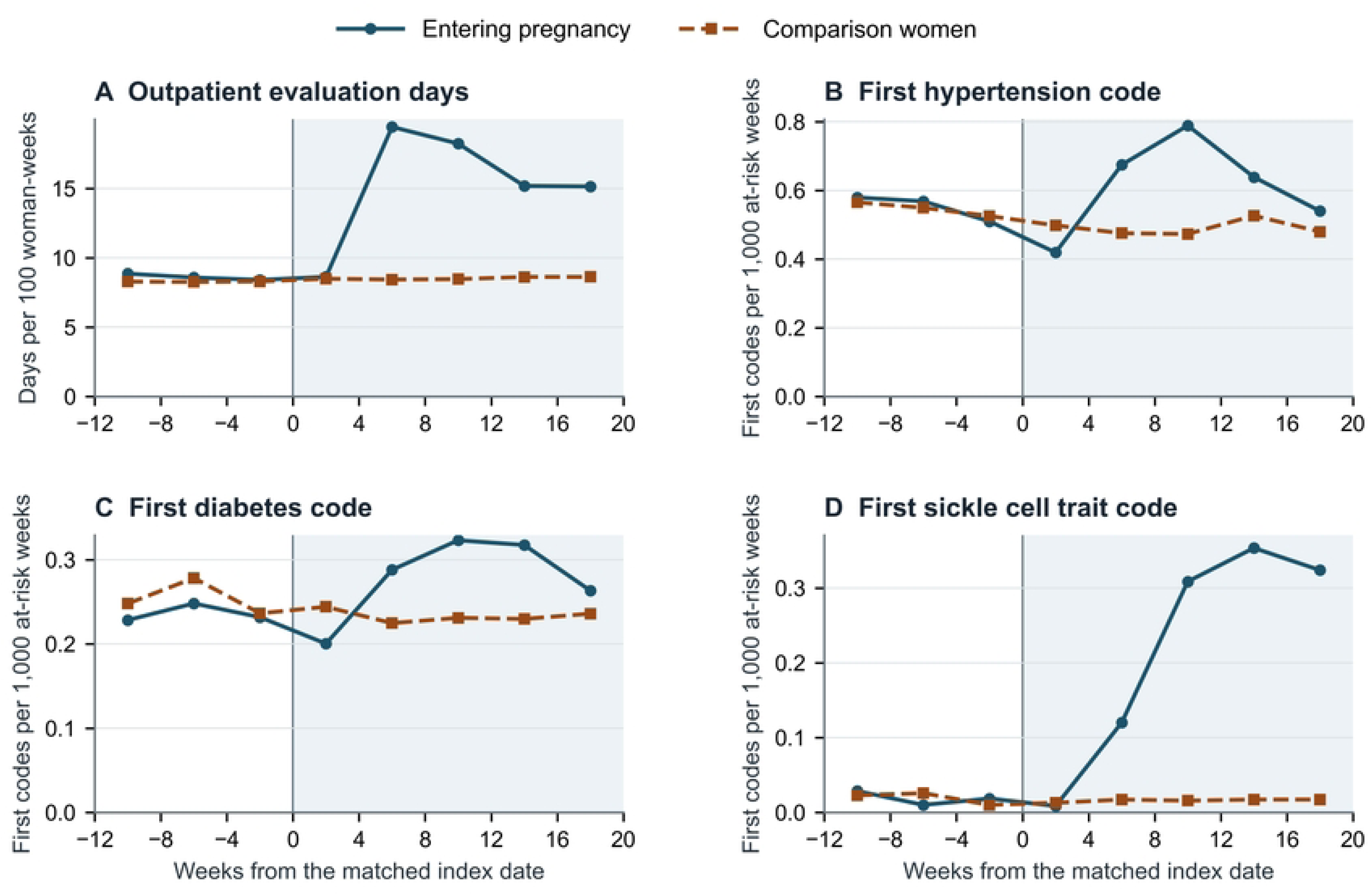
Billed outpatient evaluations and first diagnosis records around pregnancy onset. Points summarize four-week intervals. Week zero is the estimated gestational start for women entering pregnancy and the same calendar date for their matched comparison women. Diagnosis panels use time at risk among women in pairs without prior condition records in the lookback.

Before the index date, first hypertension records were nearly equally frequent in the two groups. Before 20 weeks of gestation, 12.1 per 1,000 women entering pregnancy had a first hypertension record compared with 9.7 per 1,000 comparison women. The difference was 2.36 per 1,000 (95% confidence interval 1.63 to 3.08). The difference for diabetes was smaller at 0.92 per 1,000 (0.44 to 1.41). First sickle cell trait records were also more frequent among women entering pregnancy, with a difference of 4.12 per 1,000 (3.80 to 4.44). Differences for all three conditions were already visible by 12 weeks and were statistically significant (Table 1).

**Table 1.** First diagnosis records in the matched pregnancy comparison.

| Condition | Window | Pairs | Pregnancy per 1,000 | Comparison per 1,000 | Difference per 1,000 (95% CI) |
|---|---|---|---|---|---|
| Hypertension | Before index | 159,727 | 6.61 | 6.54 | 0.07 (−0.49 to 0.63) |
|  | First 12 weeks | 157,637 | 7.47 | 5.74 | 1.73 (1.17 to 2.30) |
|  | Before 20 weeks | 157,637 | 12.09 | 9.73 | 2.36 (1.63 to 3.08) |
| Diabetes | Before index | 166,472 | 2.83 | 3.05 | −0.22 (−0.58 to 0.15) |
|  | First 12 weeks | 165,495 | 3.24 | 2.79 | 0.46 (0.09 to 0.83) |
|  | Before 20 weeks | 165,495 | 5.55 | 4.63 | 0.92 (0.44 to 1.41) |
| Sickle cell trait | Before index | 175,039 | 0.23 | 0.23 | −0.01 (−0.11 to 0.10) |
|  | First 12 weeks | 174,958 | 1.74 | 0.18 | 1.56 (1.35 to 1.77) |
|  | Before 20 weeks | 174,958 | 4.44 | 0.32 | 4.12 (3.80 to 4.44) |

Together, Fig 1 and Table 1 indicate that once a woman becomes pregnant, her healthcare contact increases. This increase leads to the diagnosis of serious diseases that previously existed but were untreated. This provides important evidence that pregnancy serves as a vital diagnostic catch-up window for diseases unrelated to birth and pregnancy.

### Larger increases among women with managed Medicaid

The payer comparison included 63,183 commercial and 107,515 managed Medicaid pairs; 4,807 pairs had other, multiple, or unknown delivery payer and were excluded from analysis. Pregnancy-group women with managed Medicaid had fewer billed outpatient evaluation days during the 365-day history period than those with commercial insurance (4.21 versus 4.86), and were more likely to have no billed evaluation during that period (22.7% versus 11.3%). Contact levels for both members of each payer group are shown in Table 2.

**Table 2.** Billed outpatient evaluation days by payer at delivery discharge.

| **Measure** | **Commercial** |  | **Managed Medicaid** |  |
|---|---|---|---|---|
|  | **Pregnancy** | **Comparison** | **Pregnancy** | **Comparison** |
| Women, n | 63,183 | 63,183 | 107,515 | 107,515 |
| Mean days, 365-day history period | 4.86 | 4.48 | 4.21 | 4.16 |
| No evaluation during history period, % | 11.3 | 13.8 | 22.7 | 23.7 |
| Mean days, 12 weeks before index | 1.18 | 1.06 | 0.96 | 0.96 |
| No evaluation before index, % | 44.4 | 48.1 | 55.2 | 55.5 |
| Mean days, first 12 weeks after index | 1.80 | 1.10 | 1.89 | 0.97 |
| Mean within-woman change | 0.63 | 0.04 | 0.94 | 0.01 |
| Pregnancy-minus-comparison difference in changes (95% CI) | 0.58 (0.57 to 0.60) |  | 0.93 (0.91 to 0.94) |  |
| Managed Medicaid minus commercial difference in changes (95% CI) | 0.34 (0.32 to 0.37) |  |  |  |

These findings indicate that women with managed Medicaid are much less likely than women with commercial insurance to have routine medical encounters before pregnancy, and thus when they become pregnant, their share of encounters increases, leading to a higher likelihood of diagnosis. During the 12-week periods immediately before and after index, mean evaluation days increased from 0.96 to 1.89 among pregnancy-group women with managed Medicaid, compared with 1.18 to 1.80 among those with commercial insurance. These increases were 0.94 and 0.63 days per woman, respectively, while changes among matched comparison women were small (Table 2; Fig 2A). After subtracting the comparison-group changes, the increases were 0.93 days for managed Medicaid and 0.58 for commercial insurance. The direct payer difference was 0.34 days per woman (95% confidence interval 0.32 to 0.37).

**Fig 2.**
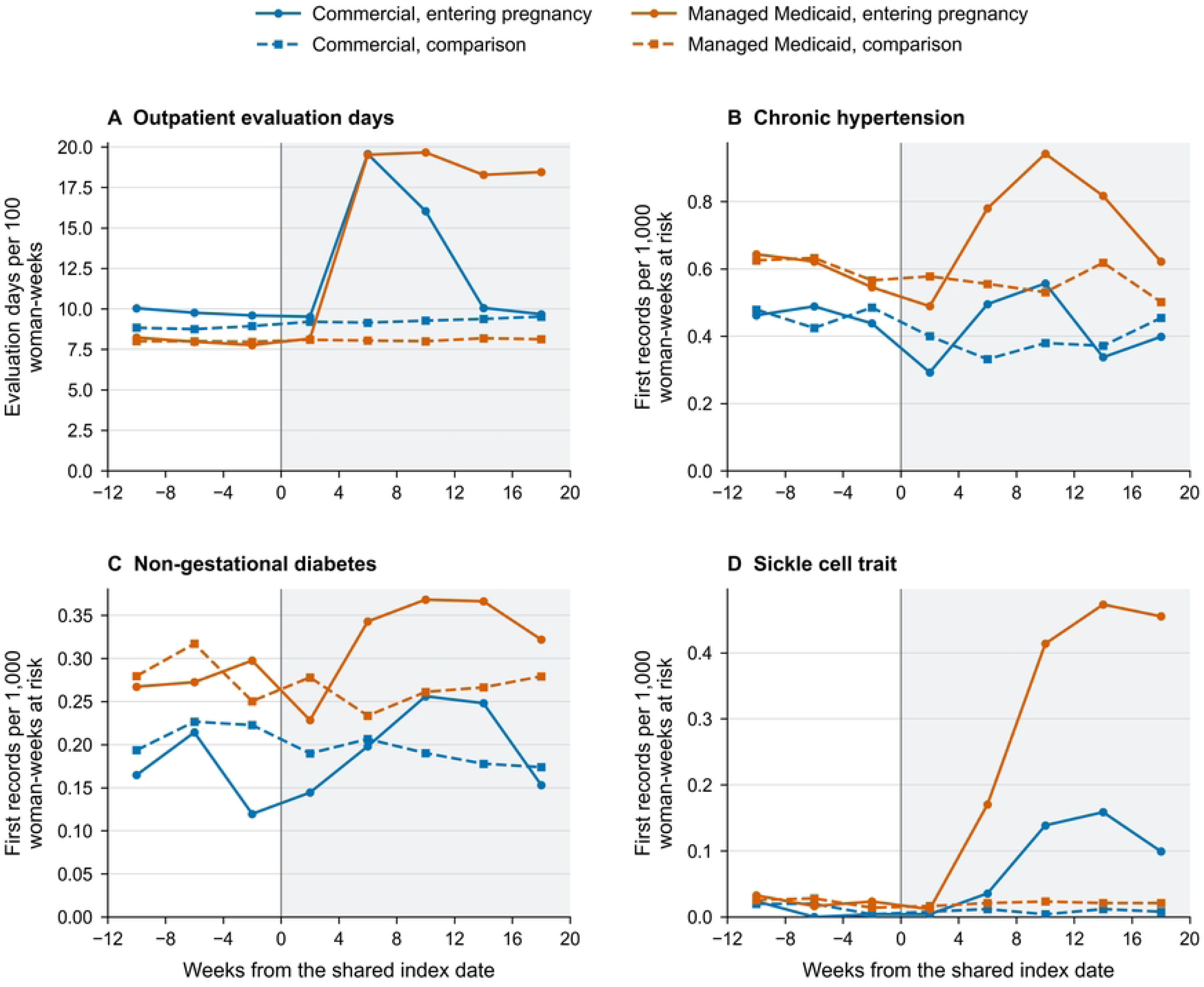
Timing of billed outpatient contact and first condition records by payer at delivery. Panel A shows outpatient evaluation days per 100 woman-weeks; panels B-D show first records of chronic hypertension, non-gestational diabetes, and sickle cell trait per 1,000 woman-weeks at risk. Points summarize four-week intervals. Week zero is the pregnancy-group woman’s estimated gestational start and the same calendar date for her matched comparison woman. Pairs are classified by the pregnancy-group woman’s medical enrollment payer at delivery discharge. Diagnosis panels exclude pairs with a condition record during the 365-day history period; each woman contributes time through her first record or the end of follow-up. These descriptive rates use the primary code definitions and do not require a billed evaluation on the diagnosis date.

Fig 2 shows that women on managed Medicaid are already more likely than women on commercial insurance to have one of the chronic traits we examine. The figure also shows that after pregnancy onset, these women see a larger increase in diagnosed conditions than women on commercial insurance. Together, these findings indicate that women on Medicaid who might have more difficulty accessing treatment see a larger spike in diagnoses during pregnancy because some of their conditions are undiagnosed prior to pregnancy onset.

Before 20 weeks, pregnancy-minus-comparison differences per 1,000 women were larger for managed Medicaid than commercial insurance for hypertension (3.35 versus 0.57), diabetes (1.26 versus 0.25), and sickle cell trait (5.65 versus 1.57). The direct payer contrasts were 2.78 (95% confidence interval 1.34 to 4.22), 1.02 (0.05 to 1.98), and 4.08 (3.49 to 4.67), respectively. Fig 2B-D shows the timing of these first records. All four principal payer contrasts had Holm-adjusted p-values below 0.05.

### Continued medical treatment

Among women with a first qualifying condition record before 20 weeks of pregnancy, we examined repeat documentation from discharge through the first anniversary of delivery, requiring continuous medical enrollment throughout this period (Fig 3A). Hypertension was recorded again in 37.9% of women with commercial insurance at delivery and 41.6% of those with managed Medicaid. The corresponding proportions were 33.9% and 37.4% for non-gestational diabetes and 12.3% and 21.0% for sickle cell trait. Importantly, these are all people who had no record of these conditions prior to pregnancy, indicating ongoing care after birth.

**Fig 3.**
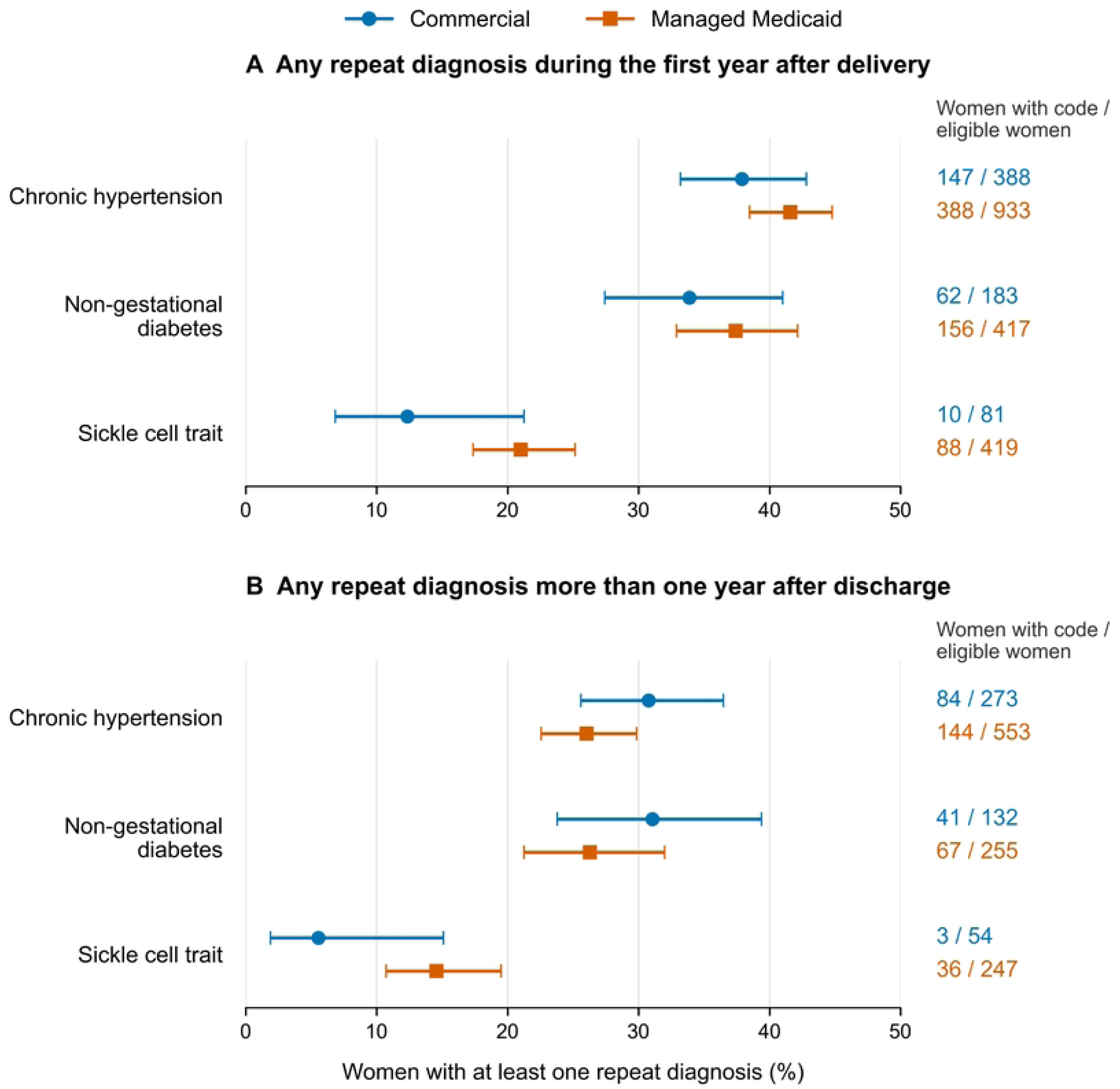
Repeat condition records after delivery, by payer at delivery. Women had a first qualifying condition record before 20 weeks of pregnancy. Panel A covers the day after discharge through the first delivery anniversary and requires continuous medical enrollment from discharge through that anniversary. Panel B covers the day after the first discharge anniversary through December 31, 2024, requiring continuous medical enrollment from discharge through that date and more than 90 days of available follow-up beyond the anniversary. Available follow-up in panel B ranged from 91 to 363 days. Eligible samples differ between panels. Points show the percentage with any repeat diagnosis record, bars show Wilson 95% confidence intervals, and labels give the number with a repeat record over the eligible sample size.

For the later follow-up analysis, we required more than 90 days (approximately three months) of available observation after the first discharge anniversary and continuous medical enrollment from discharge through December 31, 2024 (Fig 3B). This requirement excluded very short observation windows; repeat records were counted throughout each woman’s available period, which ranged from 91 to 363 days. Hypertension was recorded again in 30.8% of women with commercial insurance and 26.0% of those with managed Medicaid. The corresponding proportions were 31.1% and 26.3% for non-gestational diabetes and 5.6% and 14.6% for sickle cell trait. In both panels, the 95% confidence intervals for all payer differences included zero. Because the panels use different eligible samples and observation periods, their percentages do not establish a change in care over time. Both analyses indicate that for many women, diagnoses identified during pregnancy represent health conditions requiring treatment that continues even after the early postpartum period.

## Discussion

Many women lack easy access to regular checkups and medical procedures. For these women, pregnancy offers a fundamental change in how they interact with healthcare services. Women, especially those on Medicaid, see a large shift in their healthcare utilization once they enter pregnancy. Once these women enter the healthcare system, they undergo tests for various conditions related to pregnancy. These tests often reveal pre-existing conditions that were either undiagnosed or untreated. This paper examined three such conditions as examples to show that pregnancy offers a substantial period of diagnostic catch-up for women.

Women had more billed outpatient contact when they entered pregnancy, and more chronic conditions entered the claims record. The increase was modest for hypertension and diabetes even when we used the diagnostic codes related to non-pregnancy-specific conditions. When we included pregnancy-specific diagnosis codes for hypertension and diabetes, the increase was much larger. We focused on these non-pregnancy conditions because every mother will be tested for them in early screening, as both conditions affect pregnancy. Sickle cell trait also sees a moderate increase in diagnosis and provides clearer evidence that pregnancy brings existing conditions into view.

The sickle cell trait findings should also be considered in the context of racial and socioeconomic inequities in access to care. The trait’s higher prevalence among Black Americans and documented gaps in awareness make the first pregnancy record potentially meaningful for counseling and continuity of care [8,9]. A national survey of newborn screening programs found variation in notification and access to prior trait results [14]. Prenatal testing and discussion may therefore identify carrier status for the first time or recover information that was not communicated or retained. Realizing this opportunity requires addressing the financial, transportation, childcare, and work-related constraints that can limit care [10,11].

Our analyses did not estimate differences by race or socioeconomic status outside of what can be gleaned from payer type, and cannot establish that pregnancy reduced inequities. However, our findings indicate that inequalities in healthcare access prior to pregnancy might be a major driver of diagnoses entering the medical record around the time of pregnancy onset. Studies linking claims, screening results, and patients’ knowledge could test whether pregnancy closes these gaps and whether information is carried into care after birth.

The results were larger for women with managed Medicaid than for women with commercial insurance. Women with managed Medicaid at delivery had less billed outpatient contact before pregnancy and larger pregnancy-associated increases in contact than women with commercial coverage. Differences in first hypertension and sickle cell trait records were also larger. These patterns are consistent with pregnancy offering opportunities to document previously unrecorded conditions.

We see that, for around 30-40 percent of women with a first diagnosis in early pregnancy, treatment continues even more than a year after the birth of the child. This leads us to conclude that, for these women, the diagnosis captured a previously unknown trait for which they now receive ongoing medical care.

For many women who face limited access to healthcare services, the period of pregnancy and childbirth offers one of the most robust interactions with the healthcare system they will have in their lives. This period also puts tremendous strain on their bodies and leads to many tests. Many of these tests can reveal previously unknown or untreated conditions that require ongoing care. A woman’s medical records thus contain both diseases newly caused by the stress of pregnancy and pre-existing diseases newly detected.

For medical researchers and health professionals, it is thus important to distinguish between these possibilities as they appear in the medical record at the same time. A woman might have signs of glucose imbalance when she presents with pregnancy; this could be an imbalance driven by the pregnancy or the result of a previously undiagnosed underlying condition. This distinction matters because it leads to different treatment paths going forward. This is especially important for women on Medicaid, many of whom might lose access to their health insurance after 12 months. For all mothers who present with existing conditions at pregnancy onset, it is vital to establish postpartum care plans to continue treatment after discharge. This continued treatment can lead to a healthier life for women whose conditions were previously untreated, and the diagnostic catch-up period of pregnancy can thus be a vital part of women’s and maternal health.

## Data Availability

Data and replication materials are available at the Kansas Population Center Whelp Lab Github. https://github.com/KansasPopulationCenter/WHELP-Lab/tree/main/working-papers/pregnancy-diagnostic-catch-up

## Supporting information

**S1 Appendix. Supplementary exhibits, aggregate results, and analysis code.** Contains reproductions of Tables 1–2 and Figs 1–3 with captions, aggregate data underlying those exhibits, data dictionaries, a file-to-output guide, and analysis and figure preparation code. Licensed participant-level data are not included.

## References

1. US Preventive Services Task Force; Barry MJ, Nicholson WK, Silverstein M, Cabana MD, Chelmow D, et al. Screening for hypertensive disorders of pregnancy: US Preventive Services Task Force final recommendation statement. JAMA. 2023;330(11):1074–1082. doi: 10.1001/jama.2023.16991.

2. ElSayed NA, Aleppo G, Aroda VR, Bannuru RR, Brown FM, Bruemmer D, et al. 2. Classification and diagnosis of diabetes: Standards of Care in Diabetes—2023. Diabetes Care. 2023;46(Suppl 1):S19–S40. doi: 10.2337/dc23-S002.

3. ElSayed NA, Aleppo G, Aroda VR, Bannuru RR, Brown FM, Bruemmer D, et al. Erratum. 2. Classification and diagnosis of diabetes: Standards of Care in Diabetes—2023. Diabetes Care 2023;46(Suppl. 1):S19–S40. Diabetes Care. 2023;46(5):1106. doi: 10.2337/dc23-er05.

4. ElSayed NA, Aleppo G, Aroda VR, Bannuru RR, Brown FM, Bruemmer D, et al. Addendum. 2. Classification and diagnosis of diabetes: Standards of Care in Diabetes—2023. Diabetes Care 2023;46(Suppl. 1):S19–S40. Diabetes Care. 2023;46(9):1715. doi: 10.2337/dc23-ad08.

5. American College of Obstetricians and Gynecologists. Hemoglobinopathies in pregnancy [Internet]. 2022 Aug [cited 2026 Nov 9]. Available from: https://www.acog.org/clinical/clinical-guidance/practice-advisory/articles/2022/08/hemoglobinopathies-in-pregnancy

6. Baicker K, Taubman SL, Allen HL, Bernstein M, Gruber JH, Newhouse JP, et al. The Oregon experiment—effects of Medicaid on clinical outcomes. N Engl J Med. 2013;368(18):1713–1722. doi: 10.1056/NEJMsa1212321.

7. Patchen L, Myers MC, Budd SG, Lyu D, Fernandez SJ, Auguste TC. Prenatal remote blood pressure monitoring and detection of chronic hypertension. O G Open. 2026;3(5):e211. doi: 10.1097/OG9.0000000000000211.

8. Ojodu J, Hulihan MM, Pope SN, Grant AM; Centers for Disease Control and Prevention. Incidence of sickle cell trait—United States, 2010. MMWR Morb Mortal Wkly Rep. 2014;63(49):1155–1158.

9. Harrison SE, Walcott CM, Warner TD. Knowledge and awareness of sickle cell trait among young African American adults. West J Nurs Res. 2017;39(9):1222–1239. doi: 10.1177/0193945916665089.

10. Thompson TM, Young YY, Bass TM, Baker S, Njoku O, Norwood J, et al. Racism runs through it: examining the sexual and reproductive health experience of Black women in the South. Health Aff (Millwood). 2022;41(2):195–202. doi: 10.1377/hlthaff.2021.01422.

11. Adler A, Biggs MA, Kaller S, Schroeder R, Ralph L. Changes in the frequency and type of barriers to reproductive health care between 2017 and 2021. JAMA Netw Open. 2023;6(4):e237461. doi: 10.1001/jamanetworkopen.2023.7461.

12. Moll K, Wong HL, Fingar K, Hobbi S, Sheng M, Burrell TA, et al. Validating claims-based algorithms determining pregnancy outcomes and gestational age using a linked claims-electronic medical record database. Drug Saf. 2021;44(11):1151–1164. doi: 10.1007/s40264-021-01113-8.

13. Gourevitch RA, Natwick T, Chaisson CE, Weiseth A, Shah NT. Variation in guideline-based prenatal care in a commercially insured population. Am J Obstet Gynecol. 2022;226(3):413.e1-413.e19. doi: 10.1016/j.ajog.2021.09.038.

14. Scott JL, Christian J, Plazas Montana M, Miller YM, Naik RP. Variability in notification of positive newborn screening results for sickle cell trait across the United States. Adv Hematol. 2024;2024:3854629. doi: 10.1155/ah/3854629.

